# Risk mitigation and health communication: women’s experiences and preferences in pregnancies with high body mass index

**DOI:** 10.64898/2026.11.20.26368335

**Authors:** Tabitha Wishlade, Jennifer Jardine, Pedra Rabiee, Jamine X Kiley, Catherine Barlow, Catherine E Aiken

**Affiliations:** Department of Obstetrics and Gynaecology, University of Cambridge, Box 223, The Rosie Hospital and NIHR Cambridge Biomedical Research Centre, Cambridge CB2 0SW, United Kingdom; Department of Obstetrics and Gynaecology, Cambridge University and Royal Papworth Hospitals NHS Foundation Trust, Cambridge, CB2 0QQ, United Kingdom

## Abstract

**Background:** An increasing number of women in the UK enter into pregnancy with high BMI (BMI ≥30kg/m^2^). On a population level, higher BMI is associated with an increased risk of pregnancy complications. However, many women experience communication from healthcare professionals about BMI-related risk as unsatisfactory. We aimed to explore how these dicussions could be made more acceptable by understanding how pregnant women with high BMI perceive, experience, and receive communication about their health and risk mitigation.

**Methods:** We recruited women with a booking BMI ≥30kg/m^2^ from an antenatal ward at a tertiary maternity unit in the UK. Individual interviews were audio-recorded and transcribed. Data was analysed according to practical thematic analysis using qualitative analysis software.

**Results:** Our analysis (n=15 participants) revealed that BMI-related risk discourse resulted in a range of negative emotions, which aligned to four major themes: (i) BMI is unhelpful as a metric of health, (ii) ‘healthy pregnancy’ is not determined by BMI, (iii) BMI-related risk is emotionally burdensome, and (iv) desire to personalise BMI-related risk. Pregnant women with high BMI did not perceive themselves or their pregnancies as unhealthy and did not necessarily desire additional support or investigations in pregnancy. Women reported that maternity staff emphasised BMI-related risk but gave little information on (i) what risk means for the individual, (ii) how to reduce BMI-related risk, and (iii) why interventions are offered, and (iv) how they can be declined.

**Conclusion:** Risk mitigation and health communication may be more acceptable to pregnant women with high BMI when it acknowledges the uncertainties and limitations of BMI, prioritises outcomes and gives information that matters to women with high BMI, and avoids weight-related stigma.

## Introduction

In the UK, more than 20% of women commence pregnancy with high body mass index (BMI) ≥30kg/m^2^(1). Risk communication is an important facet of maternity care for women with high BMI as at population level they are at greater risk of outcomes such as gestational diabetes(2), pre-eclampsia, and small for gestational age(3). However women with BMI ≥30kg/m^2^ often perceive communication around pregnancy risks as unsatisfactory.

Pregnant women with high BMI describe excessive focus on their weight and pregnancy complications(4),(5),(6),(7), causing negative emotional reactions(4). This may be particularly the case when no practical lifestyle advice on how to reduce risk is given(4). BMI risk communication may also be experienced as stigmatising(8).

Simultaneously, women describe how discussion of raised BMI and pregnancy complications may be avoided by maternity healthcare professionals(4),(6),(7),(9),(10),(11) leaving some women without information about their risk status and why they have been offered BMI-related interventions(5),(10),(12). Th may lead to women feeling stigmatised(12) or seeking information from other sources(11). Further, even when information is given it may be conflicting or lack specificty(5),(9).

Effective risk communication underpins shared decision-making(13), which is in turn associated with improved pregnancy experiences(14),(15). We explore how pregnant women with high BMI perceive, experience, and receive communication about risk mitigation with the specific aim of identifying how communication around BMI in pregnancy could be made more acceptable.

## Ethical approval

The study received ethical approval from the Yorkshire & The Humber - Bradford Leeds Research Ethics Committee in November 2025 (ref: 25/YH/0230).

## Methods

### Patient and public involvement

Six women with experience of high BMI in pregnancy gave their input into the study design, materials, and interview schedule. Their input ensured that the approach to women was sensitive and acceptable, and their feedback helped to shape the interview topic guide.

### Design and setting

This qualitative descriptive study was undertaken at a tertiary NHS maternity unit in the East of England with >5000 births a year. As issues of weight, BMI, and pregnancy outcomes may be sensitive topics, especially when discussed in a healthcare environment, we undertook individual interviews in private settings instead of focus groups to ensure participants felt comfortable to share their experiences.

### Recruitment and sampling

Eligible participants were pregnant women of ≥18 years of age, ≥16^+0^ weeks’ gestation, with a booking BMI ≥30kg/m^2^ and who were able to communicate sufficiently in English to provide informed consent and participate in an interview.

Recruitment was undertaken on an antenatal ward between December 2025 and June 2026. A convenience sample of 15 women was recruited. Previous qualitative studies in similar contexts(9), (6), (7) demonstrated that ∼15-25 particiapnts typically allows for collection of rich data. Our data collection and analysis were undertaken concurrently, and the study team agreed the that the themes were sufficiently developed to address the study aims to conclude recruitment at 15 participants. Each woman received a £20 voucher as a token of gratitude for their time.

### Data collection

An interview guide was developed based on a literature review. The guide addressed experiences of risk communication in pregnancy; knowledge of BMI-related pregnancy risks; ideas of health and health optimisation both before and during pregnancy; and information- giving.

Interviews were conducted in private rooms in the antenatal setting, and lasted up to 45 minutes. Participant age, parity and booking BMI were recorded from hospital records. Interviews were recorded and transcribed using intelligent verbatim transcription.

### Analysis

Transcripts were analysed using qualitative data analysis software (ATLAS.ti Scientific Software Development GmbH, Berlin, Germany). Data analysis was guided by practical thematic analysis(16). All transcripts were coded by TW to ensure consistency and depth of engagement with the full dataset. Rigour was supported through regular discussion and co- construction of codes and developing themes with CA, JJ and PR, and through presentation to a multidisciplinary women’s health research group.

### Reflexivity

The research team consisted of TW (midwife and PhD candidate), CB (midwife), CA (obstetrician), JJ (obstetrician), PR (obstetrician), and JK (women’s health PhD candidate). TW was the interviewer and lead researcher, and was not involved in any of the participants’ clinical care. All but one of the team members had experience of providing maternity care to women with high BMI. We each reflected how this may have influenced our interpretations of the data, addressing our potential biases and blindspots.

## Results

15 women gave consent to the study and were interviewed. Their demographic data is presented in Table 1. Data were co-constructed into the themes and sub-themes presented in Figure 1.

**Figure 1:**
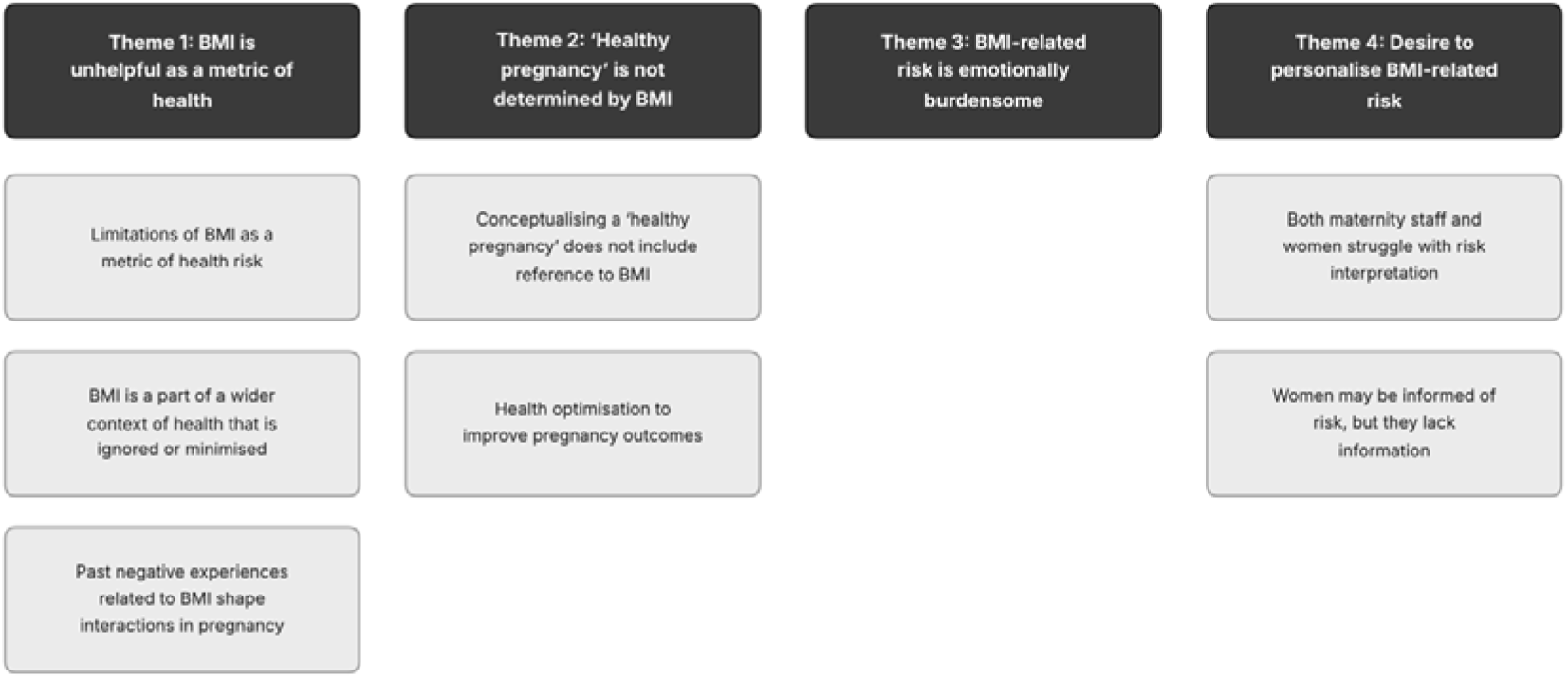
Themes and sub-themes

**Table 1:** Participant booking BMI

| **Participant** | **Booking BMI (kg/m<sup>2</sup>)** |
|---|---|
| 1 | ≥40 |
| 2 | ≥40 |
| 3 | ≥40 |
| 4 | 30-34 |
| 5 | 30-34 |
| 6 | 35-39 |
| 7 | 35-39 |
| 8 | ≥40 |
| 9 | 35-39 |
| 10 | ≥40 |
| 11 | 30-34 |
| 12 | 30-34 |
| 13 | 30-34 |
| 14 | 30-34 |
| 15 | 30-34 |

## Theme 1: BMI is unhelpful as a metric of health

### *i.* Limitations of BMI as a metric of health risk

> I generally think that someone could have a high BMI but be healthier than someone that actually has a really low BMI, be skinny, be classed as healthy when actually you don’t know what’s going on inside them fully to understand what their diet’s like, exercise is like. (012)

In differing ways, participants expressed their belief that BMI was an inadequate metric of health or health risk. Several women attributed their elevated BMI in part to their height, stating that being either tall or short pushed them into a high-risk category. One participant felt that BMI was entirely redundant as a metric of health risk for women as it was based on historical data from men.

Women discussed how BMI did not capture wider aspects of health that may impact on pregnancy. For some women, BMI had value as a broad marker of risk but needed to be interpreted in the context of other factors. Women expressed that they would welcome a risk assessment that incorporated wider social determinants of health, with specific reference to adverse life circumstances.

Participants were clear that their high BMI did not make them unhealthy, with many explicitly stating they were healthy. Some women based their assessment of health on clinical measures: ‘*I’ve always had low blood pressure’* (014). Physical activity also factored into their self-assessment: ‘*I am quite active … I’ve not felt out of breath doing things, so it’s been all right.’* (004). But the most frequently cited marker of health was their own feeling of being healthy: ‘…*Ultimately* … *I think you’d have to be led by that woman alongside other information about their health, blood pressure’* (008).

Participants communicated their health by differentiating themselves from being ‘obese’ or ‘overweight’. These words held negative health connotations that did not correlate with their lived experiences:

> I wouldn’t consider myself to be an obese person. I can cope with any form of exercise I take. I’m not on any medications. I don’t have any health conditions that would suggest that I’m impaired because of my weight or anything like that. (015)

### *ii.* BMI is a part of a wider context of health that is ignored or minimised

One woman reported that maternity staff attributed issues she faced in pregnancy to raised BMI: ‘*It BMI was very much all they talked about, any issues, pain’* (007). For example, one participant explained how her pregnancy losses were caused by an undiagnosed uterine structural anomaly and not raised BMI:

> *Do you know what my actual risk factors are? I have a bicornuate uterus, which was undiagnosed until my third pregnancy. Other than that, there is no physical specimen of evidence as to why my previous pregnancies ended in lo*sses. (015)

Women stated that their BMI not only meant they were offered extra testing in pregnancy, but were also asked to repeat sceening tests such as the glucose tolerance test even after a negative result. In some cases, this led to missed or delayed diagnoses of pregnancy complications.

Two participants further highlighted the perceived inability of clinicians to contextualise their high BMI within the broader landscape of women’s health issues including hypothyroidism, lipoedema, and PMOS. This meant lifestyle advice fell flat:

> *"Eat less and exercise more," is the advice … I do actually have a metabolic condition, I have insulin-resistant and inflammatory PCOS, which nobody seems to care about women’s health, at all. (001)*

> *They told me I needed to lose five stone … which when you’ve got a hormone problem, it’s very difficult.* (010)

### *iii.* Past negative experiences related to BMI shape interactions in pregnancy

Many women reported negative healthcare experiences related to their BMI, especially interactions where women felt they had experienced direct weight prejudice:

> ‘*It’s weight prejudice always been there, because I was always told that my infertility was due to my weight’* (001).

Many women also referenced adverse life experiences they felt were intertwined with BMI. Others had a history of health conditions linked to their BMI such as poor mental health, eating disorders, and cardiac issues. These experiences negatively shaped how participants anticipated, received, and interpreted BMI discourse within maternity services.

> *I was very much looking, expecting it weight prejudice, I suppose, in this pregnancy*. (010)

> *I wanted to establish boundaries … you’re naturally given your weight at booking*. *That can be really, really damaging for someone with a history of eating disorder, so I told my first midwives, “Please, don’t tell me.”* (015)

> *I will explain I do find this really triggering and I’d rather not, but then if people don’t respect that then I do tend to get a bit assertive, I would say. Yeah, overly assertive, yeah, and then I get really angry, and then I get really upset.* (001)

> *It [past weight prejudice] definitely does colour how I interpret interactions now with medical professionals.* (006)

## Theme 2: ‘Healthy pregnancy’ is not determined by BMI

### i. Conceptualising a ‘healthy pregnancy’ does not include reference to BMI

For most participants, healthy pregnancy was not determined by their BMI, but meant feeling healthy both physically and emotionally: ‘*I think it’s all about how you feel as a woman while you’re pregnant’* (014).

For some women a healthy pregnancy centred on *doing*, in the form of healthy eating and being active. However, participants’ views of a healthy pregnancy could be modified by their underlying health status. A respondent recently diagnosed with a chronic condition found her understanding of a healthy pregnancy had changed from that of leading a healthy lifestyle to one of communicating with her body.

> *…The first time round I’d say I just thought a healthy pregnancy was eat the right foods, stay active, do little bits, do all the sort of stuff they tell you to do, take the vitamins, whereas this time I’ve had to focus a lot more on my body, making sure it knows what I need it to do, resting when I need to but then also staying active.* (009)

Some respondents foregrounded the health of their baby as a marker of a healthy pregnancy, in some cases explicitly stating their pregnancy was healthy because the baby was well, even when they themselves were experiencing complications such as gestational diabetes or hypertension.

> *Yeah, I’d say it’s met my pregnancy is healthy, to be honest … the issues I’m facing are more with myself at the moment than baby’s, so they’ve been quite healthy and well monitored and not had any kind of growing issues massively and stuff.* (008)

Another participant took this even further, implicitly basing her assessment of a healthy pregnancy on the longer-term health of her child: ‘*I’ve talked to loads of friends who’ve had completely different pregnancies to me, and they’re all healthy children that have been delivered, so, yeah’* (015).

### ii. Health optimisation to improve pregnancy outcomes

Although most women’s conceptions of healthy pregnancy did not depend on BMI, several women stated that to optimise their health pre-pregnancy they had lost, or tried to lose, weight. Women had achieved weight loss through lifestyle changes, weight loss surgery, and use of GLP-1 agonists. In some cases, this was linked to advice from medical professionals*: ‘…Because my BMI was so over, they said that I wouldn’t be able to carry a baby and that either if the baby survived, I probably wouldn’t’* (013). Others stated they did not welcome prenatal input from clinicians as they had done what they ‘*needed and wanted to do’* (014) regarding pre-pregnancy BMI.

Generally, women were cautious about pre-pregnancy health optimisation interventions. Their caution was strongly linked to the perception that health optimisation would focus primarily on reducing their BMI.

> *I think if I was at the stage where I was not able to walk, not able to look after myself, not be able to naturally fall pregnant because of my health, then yes. But I’ve never felt at that stage in my life.* (012)

> *…Every time before that I’ve ever done any kind of health anything, “health” is a thinly veiled euphemism for “get skinny at any cost”, and I’m not interested in that.* (006)

> *Because I feel a lot of weight-related support available is very much of, "You have to have a BMI of below 30, otherwise you’re going to die at 45 of congestive heart failure,"*

> *I think is the message, really, yeah.* (001)

One respondant declined referral to a dietician in pregnancy as she understood the emphasis to be on weight loss when her focus was on the health of her baby: ‘*Maybe after pregnancy, yes, but my focus right now is just kind of keeping them the baby healthy and growing’* (008).

## Theme 3: BMI-related risk is emotionally burdensome

Some women expressed shame, anxiety, stigma and overwhelm in relation to high BMI during pregnancy. Many respondents stated they wanted and valued reassurance from maternity staff regarding their level of risk: ‘*But I think a little bit of reassurance … just go such a long way’* (010).

Women reported different strategies for navigating anxieties about being labelled ‘high risk’ owing to their BMI. These included reading information on poor outcomes to feel in control, living the healthiest lifestyle possible, or taking a more fatalistic approach to pregnancy:

> *It doesn’t matter what you do. Your body’s going to do what it wants to do. That’s exactly the attitude I feel to have because you’re just like… if you’re going to sit there and constantly worry abou*t it, then you’re just not going to enjoy anything. (002)

Frustration, exasperation and upset were common among participants, as they believed high BMI directed their maternity care to such an extent their own agency was eroded. Although there were examples of respondents declining BMI-based care, barriers to challenging the BMI care pathway - either logistical or having to fight against the weight of clinical opinion - were reported by others.

> *“Well, is it my choice?” it was, “Yes, but this is what we’re advising,” and then we’re getting second and third opinions from, you know? And it ultimately, what they say is what we want you to do. So, it felt like someone else higher up was making that decision.* (014)

> Which I didn’t feel like I had a choice … there’s no option to opt out at the midwife. *You then have to call the clinic on your own, which is hard enough, and then you have to try and cancel it. Even then, the next time you spoke to the midwife, they’d go, “Oh, you’ve not attended your diabetes test.”* (015)

## Theme 4: Desire to personalise BMI-related risk

### i. Both maternity staff and women struggle with risk interpretation

Women in the study had experienced highly variable messaging with respect to BMI and the risk of experiencing pregnancy complications, but had generally experienced communication around BMI risk as dispiriting. BMI ‘*was always a negative thing’* (014) in discussions, even in the context of significant pre-pregnancy weight loss. Some women felt that clinical staff interpreted an increased risk of poor outcomes as a certainty which was unhelpful, increased anxiety, and decreased women’s choices despite otherwise healthy pregnancies.

Women felt that many discussions with healthcare professionals lacked clarity on how to personalise their risk, although it was also noted by one participant that being given risk information at a cohort rather than individual level felt less pointed and more acceptable.

> *… it is very apparent that high BMI is a risky thing, but it’s difficult to, I suppose, navigate what that looks like for you within your pregnancy. How does that look like?* (010)

> *I was told the information but wasn’t really sure how that would really affect me and if it would affect me.* (008)

Other women wanted more specific communication of risk, including statistics.

> *You are so terrorised … This is what’s going to happen to you, not this potentially is a risk, these are the odds, these are the studies, etc. It is this is going to happen to you because of your BMI …* (001)

> *I’m still not sure, because a lot of the things say, “May,” and then I’m like, “Well, what does that mean?”* (003)

> …*But I just think there is so much that you could benefit from in terms of not just communicating what risk is but the certainty of the risk, the likelihood that it will come to pass, your evidence base for doing so.* (015)

In the absence of clear communication about their level of risk, several participants personalised and contextualised their individual risk level by using women with higher or lower BMI as frames of reference:

> *But I’ve seen bigger women be pregnant and they were okay, so I always thought I’m not that big, ….* (004)

> *My mum, she’s had four healthy pregnancies, but she’s not the healthiest person in the world. But then I’ve had friends that have struggled for months, but they’re the tiniest people you could meet.* (012)

### ii. Women may be informed of risk, but they lack information

Information on BMI-related risk was experienced as unsatisfactory for all participants, but in a variety of ways. Some women reported being given BMI-related interventions without explanation: ‘*I think it would be nice to have that explained to you a bit better, about why you need to have certain things happen to you’* (011).

Women who experienced poor pregnancy outcomes also felt that the BMI risk information they had received lacked detail, and this had contributed to how unprepared they felt when things went wrong: ‘*So, obviously, you’re recovering from giving birth, and you’re then being told that your 12-hour-old baby* is going off to neonatal care, and you’re like, “What?”*’* (002)

Further, a lack of information on reducing BMI-related risk in pregnancy was noted by many participants. This led them to surmise the only way to decrease their risk was to dramatically lose weight – something they recognised was not feasible in pregnancy, but which led to feelings of impotency.

> *I could know, also, “What can I do to decrease my risk?” because I think the answer is just, “Well, don’t be fatty fat fatty.” But we all know that that’s not a legit… unless you’re on Ozempic, which costs $3 trillion and, also, you can’t go on it in pregnancy anyway, what are some actual things I should be looking out for personally that I can do or change? I don’t know.* (006)

> *It’s not necessarily negative from a, “And here’s how you cope,” or, “And here’s how you change this,” or, “And here’s how you live with this,” or, “Here’s what you should do.” It’s just, “You’re, kind of, up a creek, sorry. Try to not have that affect you, but there’s also absolutely nothing you can do, apart from choose your pregnancy as a moment to make an exceptionally dramatic life change,” which you shouldn’t. For most people with a high BMI, you would be taking a GLP-1 … it’s, kind of, like th*e weather, like fantastic to know, but I have absolutely no control over it. So, there’s no way for me to do anything, or make any changes, or produce anything. (015)

## Discussion

We identified four over-arching themes: (i) BMI is unhelpful as a metric of health, (ii) ‘healthy pregnancy’ is not determined by BMI, (iii) BMI-related risk is emotionally burdensome, and (iv) desire to personalise BMI-related risk. These themes centred around the idea that women with high BMI did not perceive themselves or their pregnancies as unhealthy and did not necessarily desire support to achieve a healthy pregnancy beyond standard pregnancy advice. Although some women had lost weight prior to conception to optimise their health, interventions were generally viewed with suspicion if they were perceived to focus on weight loss. Talking about BMI and related risk during pregnancy was associated with a variey of negative emotions. Women reported that maternity staff emphasised BMI risk but gave little information on (i) what risk means for the individual, (ii) what women can do during pregnancy to reduce risk, and (iii) why interventions are offered, and how they can be declined.

National guidelines state women with high BMI should be provided with accurate information on BMI-related risks and how these can be minimised, along with the opportunity to discuss this information in a empowering manner(17). Conversations regarding BMI should also be sensitive and non-judgemental(18). Despite these guidelines, HCPs report lacking the confidence, knowledgeand skills needed to provide effective BMI-related discussions(19),(20),(21). Poor communication may lead to frustration(9), embarrassment(6), stigmatisation(12) and potential disengagement from care(4). Our study findings highlight areas HCPs may find helpful to consider when undertaking BMI-related health and risk conversations to achieve a positive clinical encounter.

### i. Acknowledge uncertainties and limitations

Women felt that BMI was a poor measure of adiposity and health, and mistrusted interventions they perceived as prioritising thinness over health. Specifically discussing the limitations of translating population-based epidemiological risk to individual patients(22) may be beneficial in discussing BMI in the clinical setting. Additionally, conversations with women may be more productive if HCPs acknowledge that BMI has limited value as a proxy measure of adiposity(23).

Although maternity services cannot reliably discern metabolic health in high BMI, explicitly recognising that individuals may be classified as metabolically healthy or unhealthy at any BMI(24),(25); that metabolically healthy high BMI confers no greater risk of certain poor pregnancy outcomes than overweight(26); and that in certain cases the majority of women with high BMI will have good pregnancy outcomes(27) may improve trust and decrease stigmatisation among women with high BMI — potentially improving patient engagement and health outcomes(28),(29). This matters beyond pregnancy itself because metabolic health at high BMI may not persist longitudinally(25) and therefore continued engagement could support future health promotion.

### ii. Focus on outcomes that matter to women

Patients’ responses to risk are modified by how risk outcomes are framed(30) and it is widely acknowledged that clinicians should modify discussions of risk to reflect patients’ priorities(13). Women in our study centred their ideas about healthy pregnancy around feeling well and avoiding complications for their baby. Prior research has identified the latter as a key reason pregnant women want HCPs to raise the topic of weight(31), and as motivators for engaging with health interventions(32). Conversations about health and pregnancy risk in high BMI could thus be framed to foreground these priorities. Framing conversations to be person-centred is one of the key tenets of quality maternity care(33), being associated with improved maternal and neonatal outcomes(34),(35).

### iii. Give information that matters to women

Participants sought more precise descriptions of their BMI-related risk. Study participants wanted reassurance from HCPs, which could potentially be achieved by both avoiding citing relative risk - which may increase risk perception(36) - and framing risk in positive (i.e. remaining normotensive) rather than negative (i.e. developing pre-eclampsia) terms during discussions(37).

Secondly, participants wanted their BMI risk personalised, but there are few reliable ways of stratifying women with high BMI into healthy v. at-risk groups(38). HCPs could consider highlighting other risk factors(39),(40) as points of risk contextualisation.

### iv. Acknowledge and accept

Study respondents prioritised how they felt in pregnancy as a marker of pregnancy risk, tallying with previous research(11). Self-reported health measures have shown predictive ability in other health contexts(41),(42) and may be a useful additional tool to assess risk in pregnant women next to clinicial tests(43). Following previous studies(11),(44) our participants also reported a lack of autonomy and feeling unable to decline testing. Provision of care related to BMI risk should therefore be made explicitly optional and women given the space to decline.

Finally, women in our study reported that issues arising in pregnancy were often attributed to their BMI needlessly. Reducing women’s health problems to bodyweight is a form of healthcare stigma that is present within in maternity care(45),(46). It has been suggested that such stigmatisation may be one reason why those with high BMI have worse health outcomes(47). In conceptualising and communicating BMI-related risk, HCPs should accept BMI is only one factor influencing pregnancy complications.

## Strengths and limitations

A key strength of our study is that individual interviews allowed us to explore experiences sensitively, responding to participants’ reactions as they arose. Recruits were all admitted to the antenatal ward. The inpatient setting meant that data was gathered in ‘real time’ regarding risk and its communication, and this immediacy may have allowed access to richer data. Finally, we have been able to suggest ways in which conversations about BMI, risk, and healthy pregnancy may be made more palatable for some women with high BMI.

Our study is limited due to lack of linguistic and cultural diversity within the recruits. Experiences of high BMI care may vary by ethnicity and culture, and further research is warranted in more diverse cohorts. Further, our respondents had a wide range of BMIs, which may reflect the heterogeneity in their experiences.

All participants were recruited while inpatients on the antenatal ward, and have experienced pregnancy complications. Therefore their accounts may differ from those of women with a high BMI whose antenatal care is managed entirely in outpatient settings. Future research could explore the experiences of women in specifically in community and outpatient care.

## Conclusion

Our study provides novel perspectives on the communication of risk and health outcomes for pregnant women with BMI ≥30kg/m^2^. Our study suggests that communication may be more acceptable to women with high BMI when it (i) acknowledges the limitations of BMI as a metric of adiposity and pregnancy risk, (ii) prioritises outcomes that are important to women with high BMI, and (iii) personalises risk as far as possible. This last point is an important area for future study. At present there is a distinct lack of research into defining risk groups in pregnant women with high BMI(38) despite risk personalisation being a national recommendation(13). As the number of women entering pregnancy with BMI ≥30kg/m^2^ grows globally(48), specifying BMI-related risk at an individual level would allow the provision of patient-centred care promoting optimal pregnancy outcomes for this cohort.

## Data Availability

Consent was not provided by participants for data to be made available outside of the study team.

